# Validation of Accelerometry Cut-off Points to Categorize Physical Activity Intensity Along the Pregnancy

**DOI:** 10.1101/2025.02.15.25322225

**Authors:** A. Cagiao, J. Fariñas, J. Rial-Vázquez, M. Rúa-Alonso, P.A. Díaz Brage, M. A. Giráldez, E.A. Carnero

## Abstract

**Background:** Several studies have created and validated accelerometry cut-off points to categorize intensities of physical activity (PA); however, there is not specific cut-off points in pregnant women. Pregnancy promotes specific physiological and biomechanical changes that can modify quantitively the relationship between movement and energy expenditure. The aim of this study was to develop and validate specific accelerometry cut-off points for light and moderate-vigorous PA (LPA and MVPA respectively), at each trimester of pregnancy.

**Methodology:** 31 healthy pregnant women completed the same cardiopulmonary submaximal exercise test, at each trimester of their pregnancies. Indirect calorimetry variables and accelerometry data were registered from no-dominant wrist and right hip placements at each measurement. Internal load variables (%VO_2max_, %VO2R, %HR max, %HRR and METs) were used as reference for validation and ROC analyses was performed to estimate cut-off points at each trimester and location in counts/min, MIMs and EMNO units.

**Results:** Cut-off points obtained from the ROC analysis showed a high performance with areas under curves (AUCs) above 0.90. Cut-off points tended to be higher in no-dominant wrist than in right hip location. The cut-off for MVPA on the right hip decreased after the second trimester in ENMO and MIMS units. Only slight differences between trimesters were found in cut-off points for wrist location.

**Conclusion:** Accelerometer intensity cut-off points in counts, MIMS and ENMO were validated in each trimester with internal load variables for no-dominant wrist and right hip placements.

## Introduction

Physical activity (PA) throughout pregnancy is associated with maternal health benefits and fewer newborn complications (1–3). Reducing time in sedentary behaviour and engaging in at least 20–30 minutes of moderate intensity PA on most days of the week is recommended by international guidelines (4–6). However, most of these guidelines rely on expert-committee level of evidence more than effectiveness studies (dose-response health-outcome based studies).

Accurate assessments of the amount, daily distribution and intensity of PA is essential to determine the dose-response relationship between sedentary and PA patterns and maternal-fetal health outcomes (7,8). Accelerometers (ACLs) are widely used as a feasible, objective and accurate technology to describe PA phenotype. Despite of ACLs benefits, the methodology used to collect and process the data is highly variable, which can have major impact on the interpretation of the results (9).

One of the main sources of variability in the quantification of the PA with accelerometry is related with the post-processing raw data and the selection of cut-off points to categorize the amount and intensity of PA (9). Several studies have created and validated cut-off points for general and specific populations, devices and wear location (10–13). Also, additional metrics to quantify the amount of PA from the raw data has been proposed (for example, Euclidian Norm Minus One (ENMO) units) and new cut-offs have been proposed to sustituted the traditional counts/min, which may have a significant impact in the amount of moderate-vigorous PA quantified (14).

Pregnancy promotes specific physiological changes (cardiovascular, respiratory, hormonal, and metabolic adaptations; increase in body mass, modifications of motor patterns) to meet the increased metabolic demands of the mother and foetus. These changes alter the relationship between external load and energy expenditure associated with physical activity or exercise (1,15–17). These facts preclude the use of healthy adult population cut-off points in childbering women along the pregnancy. The use of non-specific cut-off points could provide bias results due to the significant changes in body weight and associated PA energy expenditure, which is the most common paradigm utilized to validate and categorize the cut-off points for PA intensity (10).

To the best of our knowledge, there have not been any studies attempting to create and validate ACLs cut-off points for PA intensity classification in pregnant women which could impact the quantification of PA levels (18). The aim of this study was to develop and validate specific accelerometry cut-off points to categorize intensities of PA in pregnant women along pregnancy. Walking exercise paradigm was utilized to validate the cut-off points with oxygen consumption (VO_2_) as reference method. Additionally, we used heart rate (HR) as secondary validation method, and we compared results across reference methods.

## Methodology

This was a prospective follow-up study during pregnancy. Participants were recruited between April 2021 and September 2022 to perform 3 cardiopulmonary submaximal exercise tests, one at each trimester of their pregnancies. They were recruited at their reference health care center during their first visit for pregnancy status, (always before 12 weeks of gestation).

At the beginning of each assessment, clinical history and risk factors were evaluated by obstetricians and midwifes’ participants, who authorized them to perform moderate exercise. Women who were taking medication that could alter metabolism, who had diabetes prior to pregnancy, severe anemia, or uncontrolled thyroid disease, as well as those who consumed drugs, alcohol, or tobacco, were excluded of this analysis.

All procedures included in the study were approved by the ethical committee of clinical research in Galicia (Spain) (n° 2019/2013) and an Institutional Review Board of Faculty of Sport Sciences and Physical Activity at the University of A Coruna (Spain). After the screening visit at their health care center, participants signed an informed consent form (ICF) to approve enrollment in the study. Also, signed ICF included authorization to have access to their medical records and clinical history for research purposes.

The study was composed by a screening visit, 3 one-day study visits, 3 control visits and variable number of follow-up health pregnancy visits (figure 1). All assessments were completed in the same laboratory at University of A Coruña (Spain) before 12, between 16 and 18 and between 32 and 35 gestation weeks (figure 1). During the screening visit participants were instructed about the protocol as we detailed in previous study (19).

**Figure 1.**
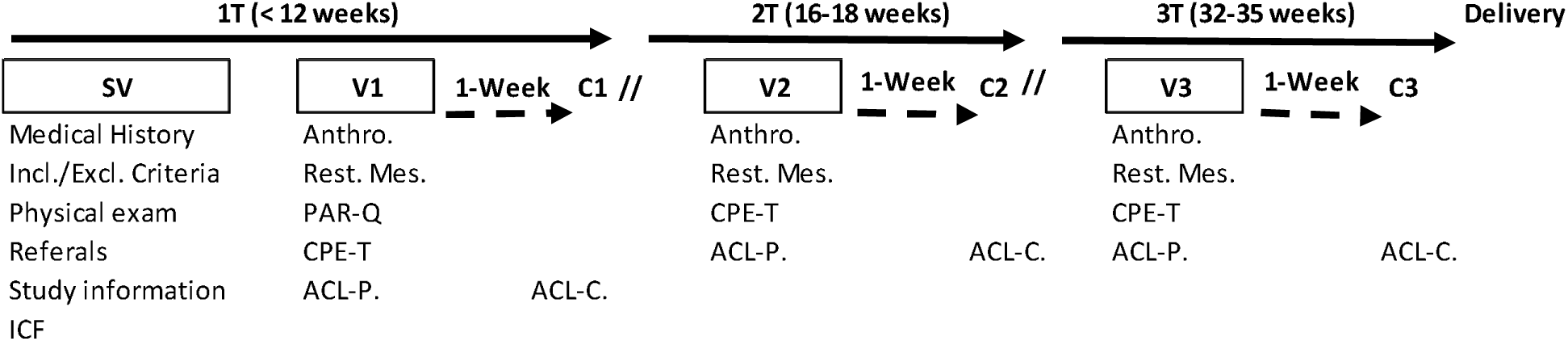
Follow-up protocol visits. 1T, first trimester; 2T, second trimester; 3T, third trimester; SV, screening visit; Incl./Excl. Criteria, inclusion and exclusion criteria; ICF, informed consent form; V1, first visit; Anthro., Anthropometry data; Rest.Mes, rest measures; PAR-Q, Physical Activity Readiness Questionnaire; CPE-T, cardiopulmonary exercise test; ACL-P, 1-week accelerometer placement; ACL-C, 1-week accelerometer control; C1, first control visit; V2, second visit; C2, second control visit; V3, third visit; C3, third control visit.

### Participants

Fifty healthy pregnant women who visited their health care centers before 12 weeks of gestation were recruited to participate in the study; however, only 31 completed the full protocol (Supplemental figure 1, Supplemental Digital Content). After the screening visit and during the follow-up visits, it was confirmed that all women had no contraindications to exercise and met the inclusion criteria. All participants delivery at term, 26 had spontaneous delivery and 5 by cesarean. All newborns were healthy, and they did not need additional care support. During the pregnancy, 3 (9.7%) women had gestational diabetes without insulin treatment needed, 4 (12.9%) had not complicated gestational hypertension and 1 (3.2%) developed type II fetal grow restriction (this condition was detected after the third assessment).

### Design

A repeated measures approach was adopted to develop cut points for each trimester of pregnancy. To create the cut points, we collected simultaneously accelerometery and internal load variables as reference for validation. Each participant exercised on a treadmill in the laboratory and wearing two ACLs (one on the non-dominant wrist and another one on the right hip; GT9X Actigraph, Pensacola, FL, USA). The oxygen consumption (VO_2_) and HR were measured every 4 minutes during 3 progressive stages of intensity using a metabolic cart (Cortex Metamax 3B, Germany), the same load per stage was repeated every trimester of pregnancy. To validate the cut points for each level of intensity, we calculated relative intensity units from estimated maximum VO_2_ (VO_2max_), maximum heart rate (HRmax) and METs (1 MET = VO_2_/3.5). VO_2max_ was estimated by simple linear regression analysis following the procedure of the American College of Sport Medicine (20). Maximum HR was calculated from Tanaka’s equation (21). To remove the metabolic effects of fetus and maternal resting metabolic rate and HR, percentages of VO_2_ reserve (%VO2R) and heart rate reserve (%HRR) were also calculated. The intensity thresholds for %VO2max, %HRmax, %VO2R and %HRR published previously (20) were followed to correlate with level of intensity by ACL as suggested by Freedson (10).

### Study visit protocol

The visits began with a brief anamnesis and the Physical Activity Readiness Questionnaire (PAR-Q) conducted by the study physician to confirm non-acute contraindications to complete the cardiopulmonary exercise test (CPE-T).Afterwards, weight and standing height were measured to the nearest 0.1 kg and 0.1 centimeters, respectively with a digital scale and stadiometer (Añó Sayol Barcelona).

ACLS were attached with a specific belt on no dominant wrist and the right hip. Configuration of ACLs was performed using the proprietary software from the manufacturer (Actilife v6.13.4) at 60 seconds epoch, 30 Hz frequency and normal filter.

### Cardiopulmonary Exercise Test (CPE-T)

A graded steady state walking exercise on a treadmill (Technogym excite med L1, Technogym, United Kingdom) was utilized as mode of exercise with simultaneous measurement of VO_2_, CO_2_ output (VCO_2_) and ventilatory variables with a portable metabolic cart (Cortex Metamax 3B) and HR with continuous electrocardiogram (EKG) (Cardio BT-100).

At the beginning of each CPE-T, environment conditions of the room were measured to confirm participants perform test under thermoneutral conditions. Gas analyzers and flow sensor were calibrated following the manufacturer recommendations with a reference gas tank and 3-L syringe, respectively. Before the start of gas collection, a basal EKG was recorded and supervised by the study physician to clear the participant to start the CPE-T. The protocol started with a 5-min resting period (3-min sitting and 2-min standing) followed by 4-min consecutive stages walking on a treadmill at 2, 4 and 6 km/h (S1, S2, S3 respectively) with 1% slope. The last minute of each stage was used to calculate all indirect calorimetry variables and accelerometry data. Actigraph ACLs registered accelerations per minute at vertical (y), horizontal (x), antero-posterior (z), and vector magnitude (VM) axis. Rated perceived exertion (RPE) was evaluated at the end of each stage by Borg scale (graded 1-10). Participants were not allowed to walk grasping the handrails. There were 2 minutes to recovery after the exercise was done, walking at 2 km/h.

### Raw data processing

We analysed raw data from Actilife software to transform gravitational units in Euclidian norm minus one (ENMO) and Monitor Independent Movement Summary units (MIMS). We used R software with GGIR and MIMS packages to transform the raw data in ENMO (22) and MIMS (23), respectively.

### Total Daily Physical Activity Assessment

Participants wore an accelerometer (GT9X Actigraph) on their non dominant wrist for 7 days after each walking test to quantify total daily physical activity (TDPA, steps/day). A daily minimum of 21 hours and at least 3 weekdays and 1 weekend day were required to consider a valid assessment for TDPA.

### Statistical analyses

Data are summarized as mean and standard deviation (SD) or standard error (SE) always variables were normally distributed. Variables non normally distributed were transformed to natural logarithm to achieve normal distribution.

Logistic regression and receiver-operating-characteristic (ROC) analyses were performed to estimate moderate-vigorous PA (MVPA) cut off points using different parameters of relative exercise intensity (%VO_2max_, %VO2R, %HR max, %HRR and METs). To calculate low intensity PA (LPA) cut-off points, direct observation was used (we used the first 5 minutes of the CPE-T as rest criterion). Performance metrics such as area under curves (AUCs), sensibility and specificity for each cut point were calculated for each parameter. ROC analyses were done to calculate cut-off points in counts, MIMS and ENMO per minute for ACL placed on no dominant wrist and right hip in each trimester of pregnancy.

All statistical procedures were performed using IBM-SPSS® 28.0 (IBM, Chicago, IL, USA) and GraphPad Prism – for Windows® (GraphPad Software Inc.).

## Results

The characteristics of the participants that completed the three assessments are described in table 1. Out of the 31 women evaluated at the beginning of the pregnancy 43.3% were normal weight, 33.3% overweight and 23.3 % obese. Body mass index (BMI) increased along the pregnancy and the weight difference between visit 1 and visit 3 (ΔWG 3T-1T) was above 15% on average, although it was highly variable. Resting HR was significantly higher during the third trimester than in the first and in the second one, which resulted in diminution of the HR reserve at the end of the pregnancy. Absolute estimated VO_2peak_ increased significantly along the pregnancy (*F*= 4.196, *P=*0.032). Total daily physical activity (steps per day) decreased significantly from the second to the third trimester (table 1).

**Table 1.**
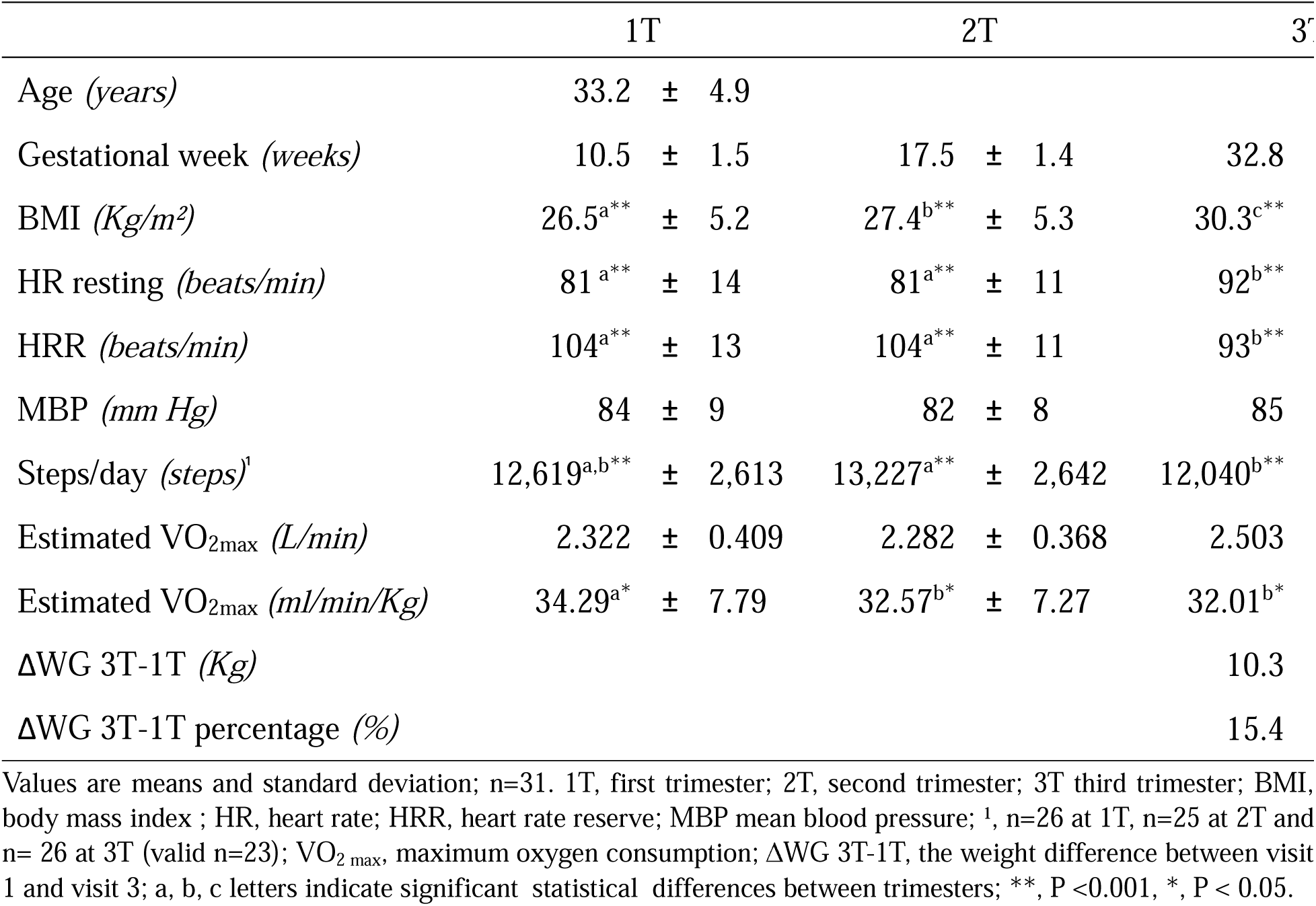
Descriptive characteristics of study participants (n=31) along the three trimesters of pregnancy.

|  | 1T | 2T | 3T |
| --- | --- | --- | --- |
| Age (*years*) | 33.2 ± 4.9 |  |  |
| Gestational week (*weeks*) | 10.5 ± 1.5 | 17.5 ± 1.4 | 32.8 |
| BMI (*Kg/m<sup>2</sup>*) | 26.5<sup>a**</sup> ± 5.2 | 27.4<sup>b**</sup> ± 5.3 | 30.3<sup>c**</sup> |
| HR resting (*beats/min*) | 81<sup>a**</sup> ± 14 | 81<sup>a**</sup> ± 11 | 92<sup>b**</sup> |
| HRR (*beats/min*) | 104<sup>a**</sup> ± 13 | 104<sup>a**</sup> ± 11 | 93<sup>b**</sup> |
| MBP (*mm Hg*) | 84 ± 9 | 82 ± 8 | 85 |
| Steps/day (*steps*)<sup>1</sup> | 12,619<sup>a,b**</sup> ± 2,613 | 13,227<sup>a**</sup> ± 2,642 | 12,040<sup>b**</sup> |
| Estimated VO<sub>2max</sub> (*L/min*) | 2.322 ± 0.409 | 2.282 ± 0.368 | 2.503 |
| Estimated VO<sub>2max</sub> (*ml/min/Kg*) | 34.29<sup>a*</sup> ± 7.79 | 32.57<sup>b*</sup> ± 7.27 | 32.01<sup>b*</sup> |
| ΔWG 3T-1T (*Kg*) |  |  | 10.3 |
| ΔWG 3T-1T percentage (%) |  |  | 15.4 |

Values are means and standard deviation; n=31. 1T, first trimester; 2T, second trimester; 3T third trimester; BMI, body mass index ; HR, heart rate; HRR, heart rate reserve; MBP mean blood pressure; <sup>1</sup>, n=26 at 1T, n=25 at 2T and n= 26 at 3T (valid n=23); VO<sub>2 max</sub>, maximum oxygen consumption; ΔWG 3T-1T, the weight difference between visit 1 and visit 3; a, b, c letters indicate significant statistical differences between trimesters; \*\*, P <0.001, \*, P < 0.05.

Accelerometer data in counts, ENMO and MIMS units increased progressively with each increment in the VO_2max%_ across different locations and trimesters (Supplemental Figures 2 and 3, Supplemental Digital Content).

Walking internal load (VO2 and HR) increased after the second trimester of pregnancy at moderate (S2, 4 km/h) and high intensity (S3,6 km/h). This increase translated in more counts, MIMs and ENMO units registered along pregnancy in S3 when ACL was worn on the wrist. Conversely, accelerometer data registered from the right hip tended to decrease significantly between trimesters in ENMO units (Supplemental Tables 1 and 2, Supplemental Digital Content).

### Cut-off points for each trimester

Estimated cut-off points for non-dominant wrist and right hip accelerometer in counts, MIMS and ENMO units for LPA and MVPA at each trimester of pregnancy are showed in tables 2-4. Parameters of relative physical activity intensity showed similar MVPA cut-off points for each ACL placement and trimester of pregnancy. Only the MET criteria provided cut-off point values significantly different than the other validation reference criteria (the furthest and lowest cut-off points in every trimester, location and cut-off units). AUCs were above 0.90 in most of the cases with high values of sensibility and specificity (tables 2-4, figures 2-3 and supplemental figures 4-7 in Supplemental Digital Content).

**Figure 2.**
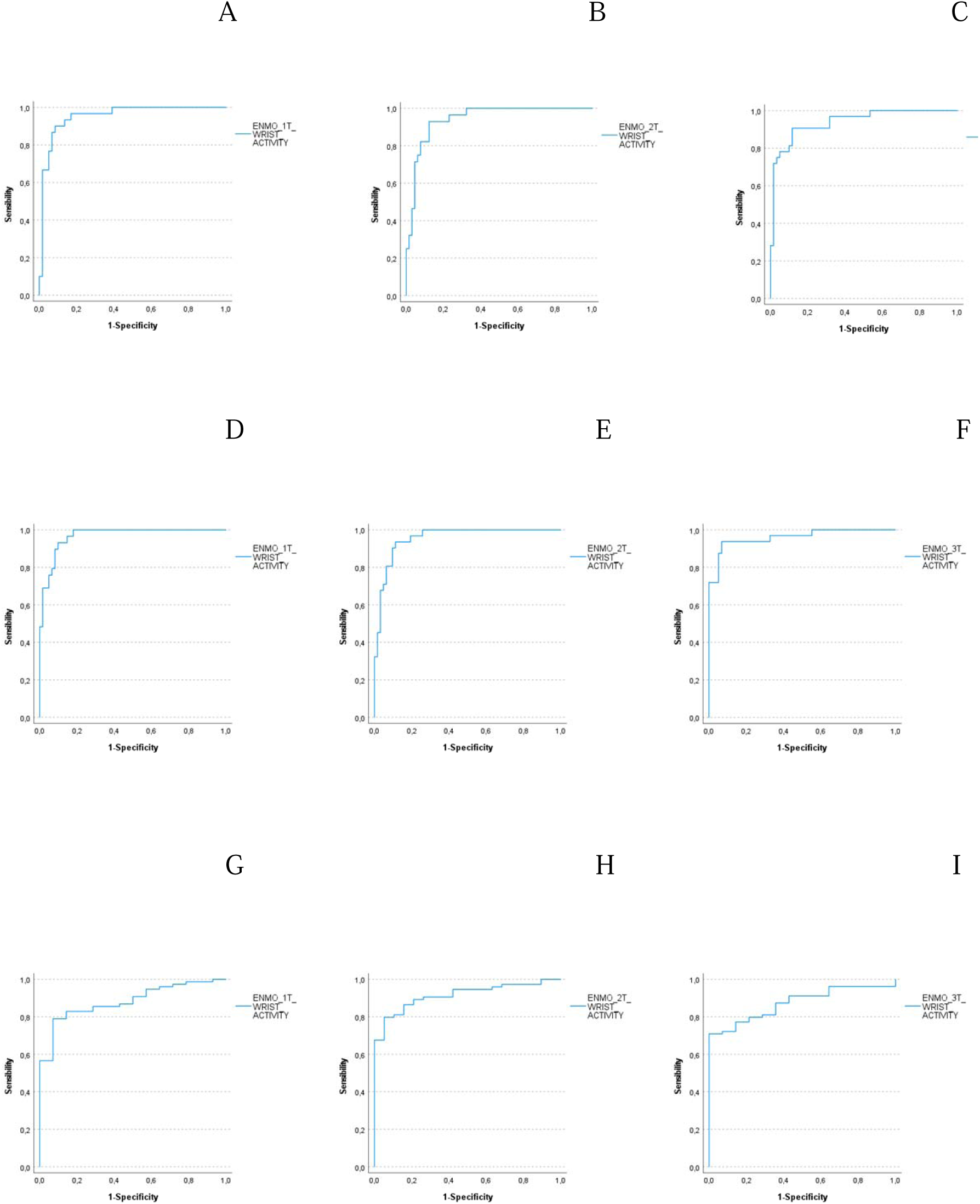
Receiver operating characteristic (ROC) curves for WRIST attachment in ENMO units and MVPA cut-off points. A, B, C are ROC curves in the first trimester (1T), the second trimester (2T) and the third trimester (3T) respectively using oxygen consumption reserve percentage (VO2R%) criterion. D, E, F are ROC curves in 1T, 2T and 3T respectively using heart rate reserve percentage (HRR%) criterion. G, H, I are ROC curves in 1T, 2T and 3T respectively using metabolic equivalent of task (MET) criterion.

**Figure 3.**
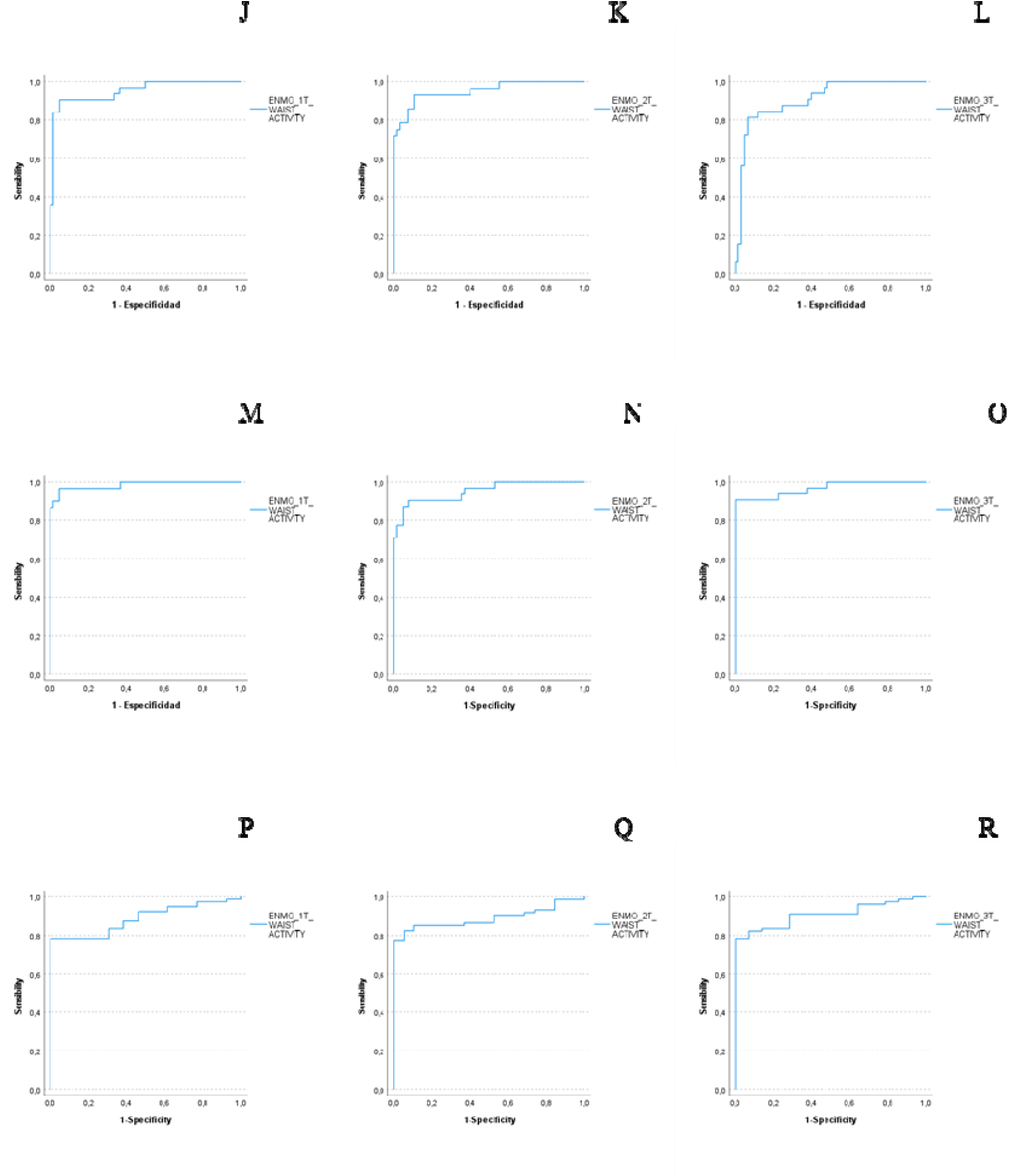
Receiver operating characteristic (ROC) curves for HIP attachment in ENMO units and MVPA cut-off points. J, K, L are ROC curves in the first trimester (1T), the second trimester (2T) and the third trimester (3T) respectively using oxygen consumption reserve percentage (VO2R%) criterion. M, N, O are ROC curves in 1T, 2T and 3T respectively using heart rate reserve percentage (HRR%) criterion. P, Q R are ROC curves in 1T, 2T and 3T respectively using metabolic equivalent of task (MET) criterion.

**Table 2.** Performance parameters of Receiver Operating Characteristics (ROC) Curves (area under ROC curve, sensitivity (%) and specificity (%)) for derived cut-off points of physical activity intensity in counts for each trimester of pregnancy.

| WRIST |  | 1T |  |  |  |  | 2T |  |  |  |  |  | 3T |  |  |  |  |  |
| --- | --- | --- | --- | --- | --- | --- | --- | --- | --- | --- | --- | --- | --- | --- | --- | --- | --- | --- |
| Reference | PA intensity | Counts | AUC | Sig. | Sens. | Spec. | PA intensity | Counts | AUC | Sig. | Sens. | Spec. | PA intensity | Counts | AUC | Sig. | Sens. | Spec. |
| %HR max | MV | 4194 | 0.85 | 0.000 | 0.85 | 0.22 | MV | 4982 | 0.89 | 0.000 | 0.85 | 0.07 | MV | 4939 | 0.85 | 0.000 | 0.73 | 0.07 |
| %VO2 max | MV | 4194 | 0.94 | 0.000 | 0.94 | 0.18 | MV | 4982 | 0.90 | 0.000 | 0.82 | 0.12 | MV | 5489 | 0.95 | 0.000 | 0.89 | 0.09 |
| %HRR | MV | 4194 | 0.93 | 0.000 | 0.97 | 0.21 | MV | 4982 | 0.95 | 0.000 | 0.94 | 0.10 | MV | 5522 | 0.97 | 0.000 | 0.94 | 0.09 |
| %VO2R | MV | 4194 | 0.93 | 0.000 | 0.93 | 0.19 | MV | 4982 | 0.92 | 0.000 | 0.89 | 0.13 | MV | 5489 | 0.95 | 0.000 | 0.94 | 0.12 |
| METs | MV | 2504 | 0.84 | 0.000 | 0.86 | 0.23 | MV | 2437 | 0.88 | 0.000 | 0.92 | 0.21 | MV | 3430 | 0.88 | 0.000 | 0.7 | 0.08 |
| Direct observation | Low | 1514 | 0.99 | 0.000 | 0.93 | 0.04 | Low | 1668 | 0.98 | 0.000 | 0.96 | 0.10 | Low | 1665 | 0.99 | 0.000 | 0.93 | 0.04 |
| HIP |  | 1T |  |  |  |  | 2T |  |  |  |  |  | 3T |  |  |  |  |  |
| Reference | PA intensity | Counts | AUC | Sig. | Sens. | Spec. | PA intensity | Counts | AUC | Sig. | Sens. | Spec. | PA intensity | Counts | AUC | Sig. | Sens. | Spec. |
| %HR max | MV | 4662 | 0.86 | 0.000 | 0.74 | 0.07 | MV | 4227 | 0.88 | 0.000 | 0.82 | 0.11 | MV | 4193 | 0.88 | 0.000 | 0.65 | 0.02 |
| %VO2 max | MV | 3804 | 0.96 | 0.000 | 0.91 | 0.12 | MV | 4104 | 0.91 | 0.000 | 0.85 | 0.14 | MV | 4239 | 0.87 | 0.000 | 0.74 | 0.13 |
| %HRR | MV | 4662 | 0.97 | 0.000 | 0.90 | 0.05 | MV | 4227 | 0.92 | 0.000 | 0.90 | 0.11 | MV | 4235 | 0.94 | 0.000 | 0.88 | 0.07 |
| %VO2R | MV | 3967 | 0.96 | 0.000 | 0.94 | 0.12 | MV | 4227 | 0.92 | 0.000 | 0.93 | 0.13 | MV | 4239 | 0.88 | 0.000 | 0.78 | 0.12 |
| METs | MV | 2414 | 0.86 | 0.000 | 0.79 | 0.08 | MV | 2468 | 0.87 | 0.000 | 0.85 | 0.07 | MV | 2640 | 0.91 | 0.000 | 0.77 | 0.00 |
| Direct observation | Low | 694 | 1.00 | 0.000 | 1.00 | 0.00 | Low | 673 | 1.00 | 0.000 | 1.00 | 0.00 | Low | 714 | 1.00 | 0.000 | 1.00 | 0.00 |

**Table 3.** Performance parameters of Receiver Operating Characteristics (ROC) Curves (area under ROC curve, sensitivity (%) and specificity (%)) for derived cut-off points of physical activity intensity in MIMS (Monitor-Independent Movement Summary units) units for each trimester of pregnancy.

| WRIST |  | 1T |  |  |  |  |  | 2T |  |  |  |  |  | 3T |  |  |  |  |
| --- | --- | --- | --- | --- | --- | --- | --- | --- | --- | --- | --- | --- | --- | --- | --- | --- | --- | --- |
| Reference | PA intensity | MIMS | AUC | Sig. | Sens. | Spec. | PA intensity | MIMS | AUC | Sig. | Sens. | Spec. | PA intensity | MIMS | AUC | Sig. | Sens. | Spec. |
| %HR max | MV | 25.19 | 0.86 | 0.000 | 0.82 | 0.14 | MV | 22.83 | 0.86 | 0.000 | 0.88 | 0.20 | MV | 25.61 | 0.84 | 0.000 | 0.71 | 0.05 |
| %VO2 max | MV | 25.04 | 0.93 | 0.000 | 0.94 | 0.12 | MV | 22.83 | 0.89 | 0.000 | 0.88 | 0.20 | MV | 26.60 | 0.92 | 0.000 | 0.86 | 0.12 |
| %HRR | MV | 25.19 | 0.94 | 0.000 | 0.97 | 0.13 | MV | 22.83 | 0.93 | 0.000 | 0.97 | 0.18 | MV | 26.86 | 0.96 | 0.000 | 0.91 | 0.07 |
| %VO2R | MV | 25.04 | 0.93 | 0.000 | 0.93 | 0.14 | MV | 22.83 | 0.90 | 0.000 | 0.93 | 0.23 | MV | 26.60 | 0.93 | 0.000 | 0.91 | 0.13 |
| METs | MV | 18.16 | 0.80 | 0.000 | 0.71 | 0.21 | MV | 20.91 | 0.85 | 0.000 | 0.70 | 0.16 | MV | 18.27 | 0.81 | 0.000 | 0.77 | 0.29 |
| Direct observation | Low | 10.21 | 1.00 | 0.000 | 0.98 | 0.02 | Low | 12.93 | 0.99 | 0.000 | 0.96 | 0.03 | Low | 10.12 | 1.00 | 0.000 | 0.99 | 0.03 |

| HIP |  | 1T |  |  |  |  |  | 2T |  |  |  |  |  | 3T |  |  |  |  |
| --- | --- | --- | --- | --- | --- | --- | --- | --- | --- | --- | --- | --- | --- | --- | --- | --- | --- | --- |
| Reference | PA intensity | MIMS | AUC | Sig. | Sens. | Spec. | PA intensity | MIMS | AUC | Sig. | Sens. | Spec. | PA intensity | MIMS | AUC | Sig. | Sens. | Spec. |
| %HR max | MV | 29.20 | 0.87 | 0.000 | 0.80 | 0.09 | MV | 30.29 | 0.91 | 0.000 | 0.82 | 0.08 | MV | 24.00 | 0.87 | 0.000 | 0.69 | 0.05 |
| %VO2 max | MV | 29.20 | 0.95 | 0.000 | 0.85 | 0.07 | MV | 25.60 | 0.93 | 0.000 | 0.85 | 0.13 | MV | 27.98 | 0.88 | 0.000 | 0.77 | 0.07 |
| %HRR | MV | 29.20 | 0.98 | 0.000 | 0.97 | 0.07 | MV | 30.69 | 0.95 | 0.000 | 0.90 | 0.05 | MV | 27.98 | 0.96 | 0.000 | 0.91 | 0.03 |
| %VO2R | MV | 29.20 | 0.95 | 0.000 | 0.90 | 0.07 | MV | 30.29 | 0.95 | 0.000 | 0.93 | 0.09 | MV | 27.98 | 0.90 | 0.000 | 0.81 | 0.07 |
| METs | MV | 18.75 | 0.89 | 0.000 | 0.79 | 0.00 | MV | 18.03 | 0.88 | 0.000 | 0.84 | 0.00 | MV | 16.74 | 0.91 | 0.000 | 0.79 | 0.00 |
| Direct observation | Low | 6.56 | 1.00 | 0.000 | 1.00 | 0.00 | Low | 6.91 | 1.00 | 0.000 | 1.00 | 0.00 | Low | 7.73 | 1.00 | 0.000 | 1.00 | 0.00 |

**Table 4.** Performance parameters of Receiver Operating Characteristics (ROC) Curves (area under ROC curve, sensitivity (%) and specificity (%)) for derived cut-off points of physical activity intensity in ENMO (Euclidian Norm Minus One) units for each trimester of pregnancy.

| WRIST |  | 1T |  |  |  |  |  | 2T |  |  |  |  |  | 3T |  |  |  |  |
| --- | --- | --- | --- | --- | --- | --- | --- | --- | --- | --- | --- | --- | --- | --- | --- | --- | --- | --- |
| Reference | PA intensity | ENMO | AUC | Sig. | Sens. | Spec. | PA intensity | ENMO | AUC | Sig. | Sens. | Spec. | PA intensity | ENMO | AUC | Sig. | Sens. | Spec. |
| %HR max | MV | 129.220 | 0.88 | 0.000 | 0.85 | 0.14 | MV | 135.290 | 0.89 | 0.000 | 0.82 | 0.15 | MV | 149.135 | 0.86 | 0.000 | 0.65 | 0.02 |
| %VO2 max | MV | 172.135 | 0.96 | 0.000 | 0.91 | 0.07 | MV | 113.560 | 0.91 | 0.000 | 0.91 | 0.20 | MV | 139.570 | 0.93 | 0.000 | 0.89 | 0.12 |
| %HRR | MV | 172.135 | 0.97 | 0.000 | 0.93 | 0.10 | MV | 135.290 | 0.96 | 0.000 | 0.94 | 0.11 | MV | 149.135 | 0.96 | 0.000 | 0.94 | 0.07 |
| %VO2R | MV | 172.135 | 0.95 | 0.000 | 0.90 | 0.09 | MV | 144.995 | 0.94 | 0.000 | 0.93 | 0.12 | MV | 146.700 | 0.94 | 0.000 | 0.91 | 0.12 |
| METs | MV | 79.285 | 0.88 | 0.000 | 0.79 | 0.07 | MV | 87.544 | 0.92 | 0.000 | 0.80 | 0.05 | MV | 93.126 | 0.87 | 0.000 | 0.71 | 0.00 |
| Direct observation | Low | 46.812 | 1.00 | 0.000 | 0.96 | 0.01 | Low | 48.048 | 0.99 | 0.000 | 0.93 | 0.08 | Low | 38.350 | 0.99 | 0.000 | 0.99 | 0.07 |
| HIP |  | 1T |  |  |  |  |  | 2T |  |  |  |  |  | 3T |  |  |  |  |
| Reference | PA intensity | ENMO | AUC | Sig. | Sens. | Spec. | PA intensity | ENMO | AUC | Sig. | Sens. | Spec. | PA intensity | ENMO | AUC | Sig. | Sens. | Spec. |
| %HR max | MV | 151.745 | 0.89 | 0.000 | 0.80 | 0.07 | MV | 130.710 | 0.90 | 0.000 | 0.82 | 0.10 | MV | 106.975 | 0.84 | 0.000 | 0.65 | 0.02 |
| %VO2 max | MV | 151.745 | 0.95 | 0.000 | 0.85 | 0.05 | MV | 130.710 | 0.92 | 0.000 | 0.79 | 0.12 | MV | 131.385 | 0.90 | 0.000 | 0.77 | 0.07 |
| %HRR | MV | 151.745 | 0.98 | 0.000 | 0.97 | 0.05 | MV | 156.965 | 0.95 | 0.000 | 0.87 | 0.05 | MV | 143.790 | 0.97 | 0.000 | 0.91 | 0.00 |
| %VO2R | MV | 151.745 | 0.95 | 0.000 | 0.90 | 0.05 | MV | 130.710 | 0.95 | 0.000 | 0.93 | 0.11 | MV | 132.425 | 0.91 | 0.000 | 0.81 | 0.07 |
| METs | MV | 81.413 | 0.89 | 0.000 | 0.79 | 0.00 | MV | 71.386 | 0.89 | 0.000 | 0.82 | 0.05 | MV | 59.794 | 0.91 | 0.000 | 0.79 | 0.00 |
| Direct observation | Low | 46.112 | 0.94 | 0.000 | 0.88 | 0.17 | Low | 35.610 | 0.97 | 0.000 | 0.88 | 0.10 | Low | 21.762 | 0.97 | 0.000 | 0.96 | 0.14 |

Discrimination of LPA was almost perfect, with the area under the ROC curve at or exceeding 0.94 (ROC curves for LPA in Supplemental figures 8-10, Supplemental Digital Content). After excluding MET criterion, the mean of the MVPA cut-off points with AUC ≥ 90 determined the cut-off points during pregnancy (table 5).

**Table 5.** Proposed cut-off points for criteria of physical activity intensity, accelerometry unit, accelerometer body attachment and trimester of pregnancy.

|  |  | 1T | 2T | 3T |
| --- | --- | --- | --- | --- |
| Wrist counts | LPA | 1514 | 1668 | 1665 |
|  | MVPA | 4194 | 4982 | 5500 |
| Hip counts | LPA | 694 | 673 | 714 |
|  | MVPA | 4144 | 4186 | 4235 |
| Wrist MIMS | LPA | 10.21 | 12.93 | 10.12 |
|  | MVPA | 25.09 | 22.83 | 26.69 |
| Hip MIMS | LPA | 6.56 | 6.91 | 7.73 |
|  | MVPA | 29.20 | 29.22 | 27.98 |
| Wrist ENMO | LPA | 46.812 | 48.048 | 38.350 |
|  | MVPA | 172.135 | 131.282 | 145.135 |
| Hip ENMO | LPA | 46.112 | 35.610 | 21.762 |
|  | MVPA | 151.745 | 137.274 | 135.867 |

LPA, light physical activity; MVPA, moderate-vigorous physical activity; 1T, first trimester; 2T second trimester; 3T, third trimester.

LPA and MVPA cut-off points tended to be higher when ACL was placed on the wrist than on the hip, except for MIMs units which MVPA cut-off points were higher on the hip. Similar tendencies during pregnancy were found for MVPA cut-off points in ENMO and MIMS units; thus, when the ACL was placed on the hip the cut-off points were significantly lower after the second trimester (tables 3,4). Moreover, MVPA cut-offs points in counts increased along pregnancy when ACL was worn on the wrist and slight variability was found when ACL was on the hip (table 2).

## Discussion

This study identified cut-off points for different ACL locations and metrics at each trimester of pregnancy. The ROC analyses discriminated cut-offs for LPA and MVPA successfully (high AUC and sensitivity). Also, following the current trend in the field of PA research (9,22,24–28); we provided the cut-off points in several metric units, such as counts, MIMs and ENMO which will permit comparations between future and previous studies.

Our study utilized several paradigms of internal load to calibrate and validated our cut-off points, which adds internal consistency to our results and allow comparisons with previous studies in non-pregnant populations. Most of the calibration studies stablish the relation between ACL counts and measured activity energy expenditure by using MET units and assuming an estimated resting expenditure of 1MET=3.5 mL/Kg/min in their calculation; moreover, the MET criterion assumes all individuals achieve the same intensity threshold at the same absolute energy expenditure (MET units). These two previous assumptions must limit the utilization of absolute METs like intensity criterion in studies including participants with heterogenous metabolic phenotypes. This limitation of the MET unit was confirmed in our study, since out of the 5 parameters of internal load that we used as criteria to categorize exercise intensity (%VO2max; %HR max; %VO2R; %HRR and MET), MET criterion performed significantly different from the others for each trimester and location. This discrepancy may be due to the increased resting energy expenditure observed along pregnancy (29,30) and the fact that absolute METs are not able to differentiate fitness level (31). Therefore, MET criterion does not appear to be a valid reference criterion to identify relative intensity of PA/exercise in pregnant women (32).

We were not able to compare the new cut-off points with others developed in pregnant women. Only a previous study has proposed PA intensity threshold for pregnant women (32); however, the authors did not provide enough statistical description and data to interpret their results with our counts/min cut-offs. When comparing our cut-off points in counts/min with other populations; overall, MVPA cut-off points (counts/min) during pregnancy are higher than cut-off points determined in non-gestational adult population when triaxial accelerometers are placed on the right hip (Santos-Lozano et al., ≤ 3208, 3209-8565 and ≥ 11,593 counts per minute for moderate, vigorous and very vigorous PA respectively; Sasaki et al., 2691-6166, 6167-9642, ≥ 9643 counts per minute for moderate, vigorous and very vigorous PA respectively) (13,33) and similar in the first trimester of pregnancy when ACL is attached to the non-dominant wrist (Montoye et al., 3941 counts per min for MVPA) (34). This suggests that the wrist must be a more stable and reliable location to place the accelerometer since it could be less prone to the walking biomechanical alterations related with pregnancy (35,36). Finally, to ensure more compliance, this placement could be preferable during later stages of pregnancy (18,22,37).

Regarding new accelerometery metrics (ENMO and MIMs), we are not able to compare our results with previous studies because few calibration studies have been performed with a GT9X ACL (the model used in this study) and the main cut-off points used in research were developed with uniaxial ACLs (10,12,38) making impossible to recalculated the new units from old uniaxial ACL data. Hildebrand et al. developed cut-off points for non-dominant wrist and right hip processing raw data in ENMO units. Our cut-off points in ENMO for MVPA are close to the moderate PA cut-off points for children defined in Hildebrand’s study (201.4 and 142.6, for wrist and hip location respectively) (22). Currently, we are not aware of validated cut-off points in MIMs units and only observational studies had used these new units (23,25,26,39). Using a GT9X ACL on the wrist, Albinaly reported 25 and 140 MIMS in 4.8 km/h and 8.8 km/h, respectively; these values were similar with the MVPA cut-off points we found in the first trimester of pregnancy (25.09 in the wrist).

To create LPA cut-off points, we used direct observation like reference method, which included a period of 3 minutes sitting and 2 minutes standing rather than a standard measure of LPA. The LPA cut-off point in the first trimester of pregnancy was very similar to the same cut-off point identified by Hildebrand et al. in ENMO units when ACL was on hip (44.8 mg, (24)).

Anatomical and physiological modifications during pregnancy could explain the fact that after second trimester of pregnancy the MVPA cut-off points on hip were lower in ENMO and MIMs units than for the first trimester. This could be explained by an increase in walking energy expenditure along the pregnancy (19,40–43); thus, the cut-off would be lower to achieve the same relative exercise intensity (VO_2max_ or HR_max_ percentage). Regarding MVPA cut-off points in counts from wrist location, they increased during pregnancy. A possible explanation for this change must be related with the weight gain during pregnancy which, can affects walking biomechanics. Thus, pregnant women must need to move more the wrist to support the same speed during pregnancy in order to maintain their impaired balance and increased stride cadence (36).

### Strengths and limitations

Physiological changes during pregnancy call for using specific accelerometer cut-off points in this population (15). This is the first study that create and validate specific accelerometry cut-off points during the three trimesters of pregnancy. Moreover, we developed intensity cut-off points in several units and locations for each trimester of pregnancy. Also, the intensity cut-off points were validated using as reference several measures of internal load during pregnancy, seeking to compare relative vs. absolute PA intensity (44).

VO_2max_ and HRmax were calculated by strapolation using direct measurements of submaximal VO_2_ and HR, this was due to difficults to pass an Institutional Review Board when maximal exercise was included in the protocol. We used only walking on a treadmill as mode of exercise and asume that can be use as a subrogate of overall ambulatory daily movement. Future research would be neccessary to cross-validate these cut-points for alternative activities and for walking under free-living conditions as derived intensity thresholds are influenced by activities chosen when performing the calibration studies (9,45).

### Conclusion

We developed specific accelerometry cut-off points to assess levels PA in pregnant women at each trimester of pregnancy for non-dominate wrist and right hip locations and for several units (counts/min, ENMO and MIMS). The stability of the derived cut-offs across the several validation reference methods/criteria (%VO2 max, %VO2R, %HR max, %HRR) may be a proof of internal consistency that needs to be confirm in cross-validation studies.

## Data Availability

All data produced in the present study are available upon reasonable request to the authors

