## Supplemental Figure 1 for "Validation of Accelerometry Cut-off Points to Categorize Physical Activity Intensity Along the Pregnancy"

**Figure 1.** Flowchart

47 pregnant women agree to participate

First trimester

39 valid measures

Second trimester

36 valid measures

Third trimester

31 valid measures

50 pregnant women recruited

3 losses:

2 miscarriages

1 give up

3abandened

5 losses:

1 preterm delivery

1 miscarriage

2 configuration fails

31 pregnant women with 3 valid measures

8 losses:

4 miscarriages

1 configuration fail

3 give up
