## Supplementary figures and images for "Validation of Accelerometry Cut-off Points to Categorize Physical Activity Intensity Along the Pregnancy"

### Supplemental Figure 2

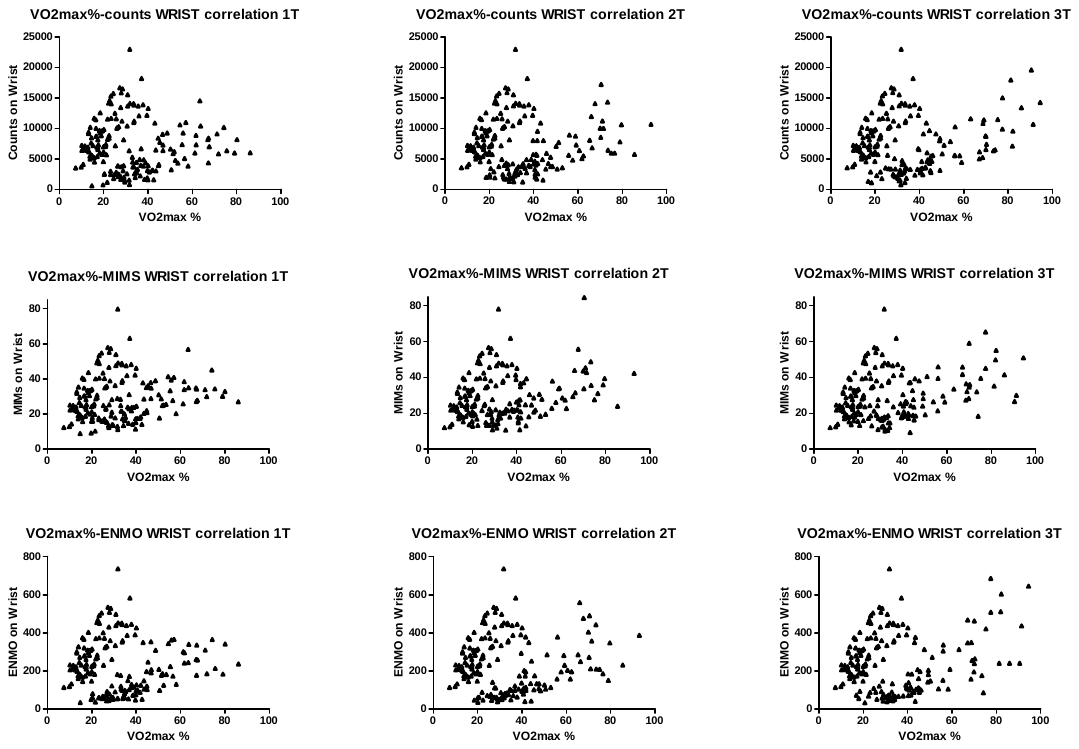

*Figure 2. VO2max % correlations when the accelerometer was placed on the wrist.*

### Supplemental Figure 3

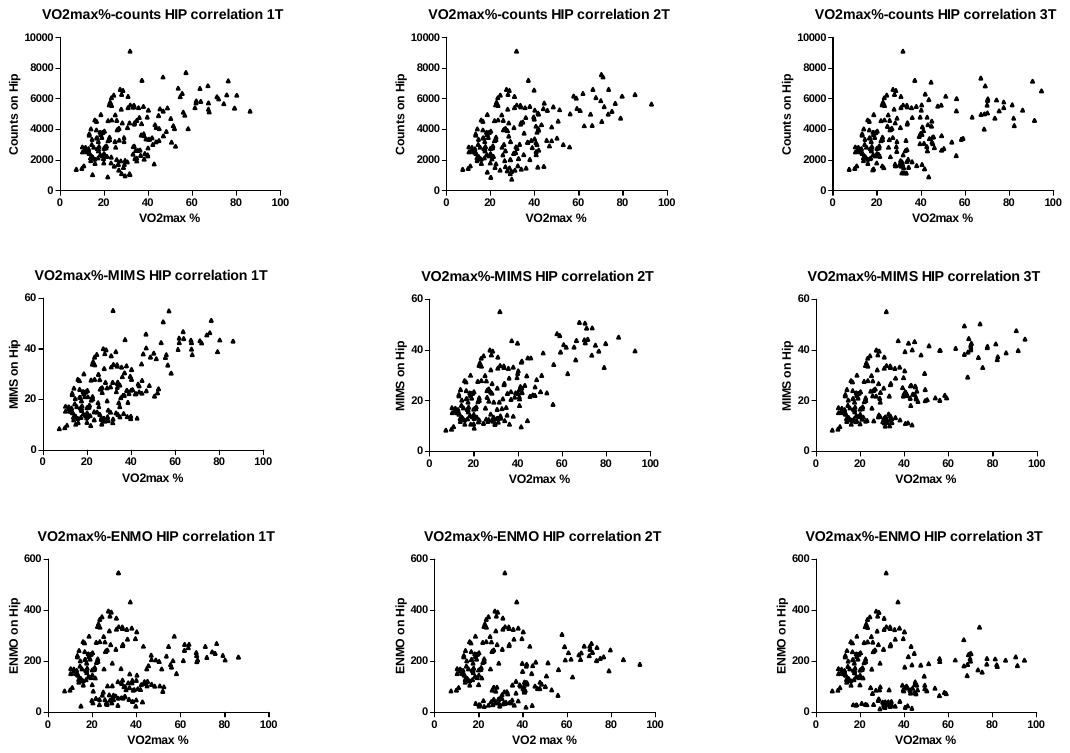

*Figure 2. VO2max % correlations when accelerometer was placed on the hip.*
