## Supplemental Figure 5 for "Validation of Accelerometry Cut-off Points to Categorize Physical Activity Intensity Along the Pregnancy"

J	K	L

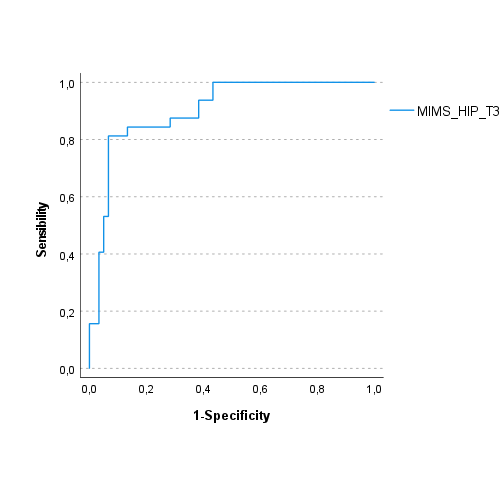

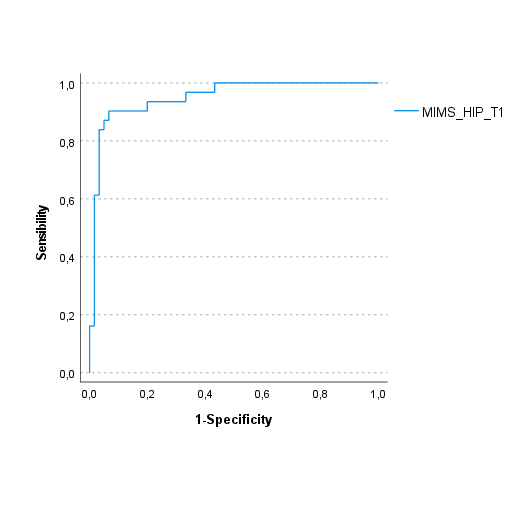

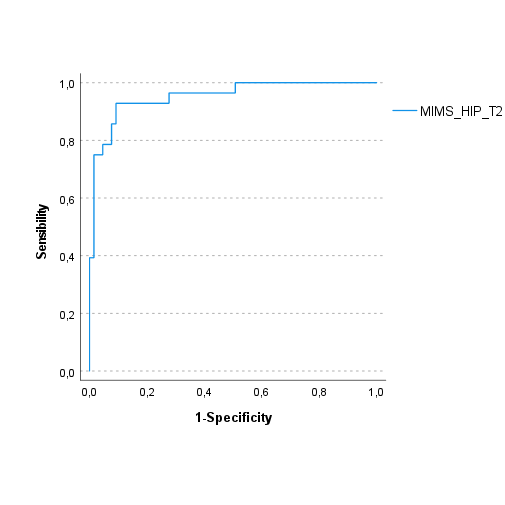

M	N	O

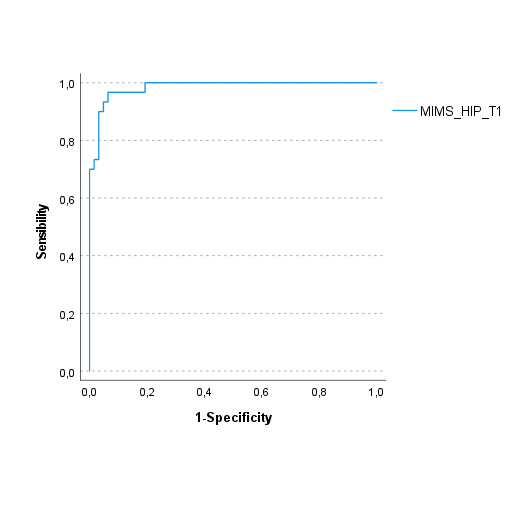

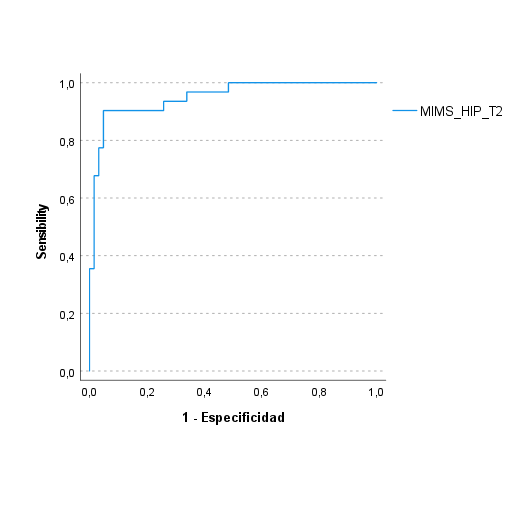

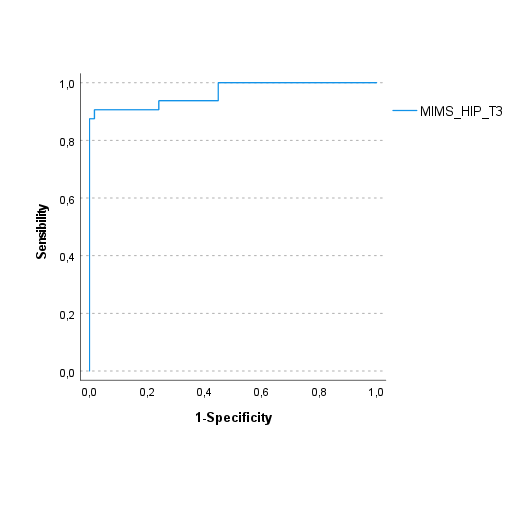

P	Q	R

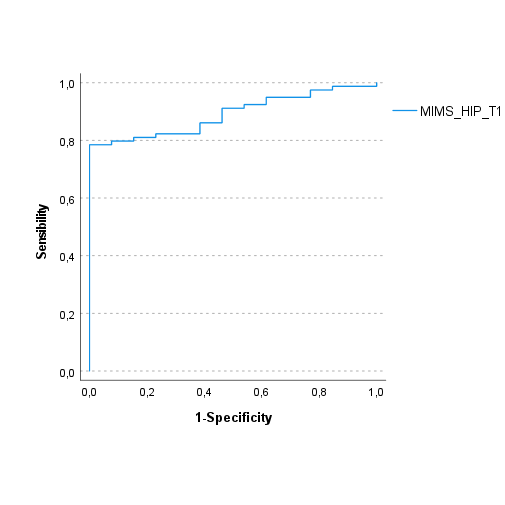

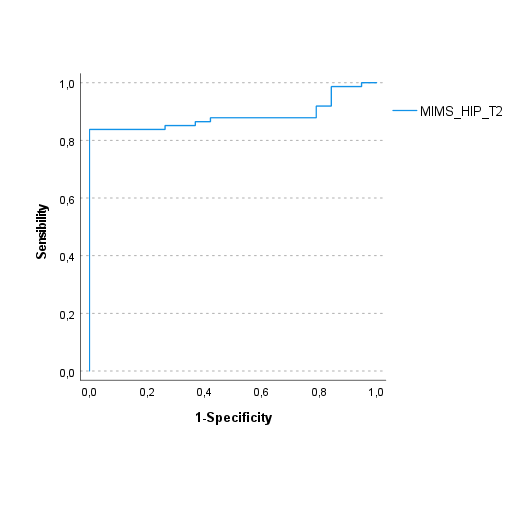

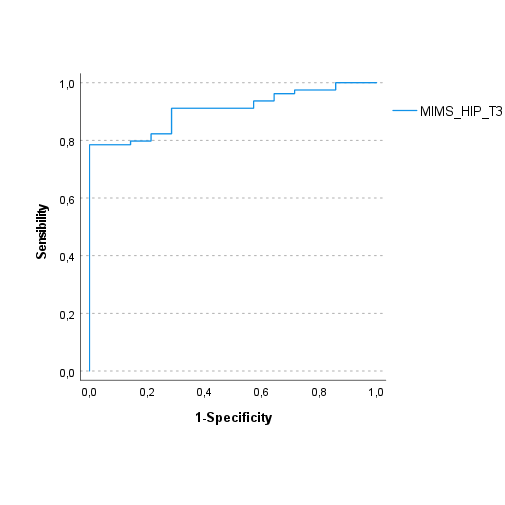

**Figure 4.** ROC curves for HIP attachment in MIMS units and MVPA cut-off points. J, K, L are ROC curves in 1T, 2T and 3T respectively using VO2R% criterion. M, N, O are ROC curves in 1T, 2T and 3T respectively using HRR% criterion. P, Q, R are ROC curves in 1T, 2T and 3T, respectively using MET criterion.
