## Supplemental Figure 6 for "Validation of Accelerometry Cut-off Points to Categorize Physical Activity Intensity Along the Pregnancy"

A	B	C

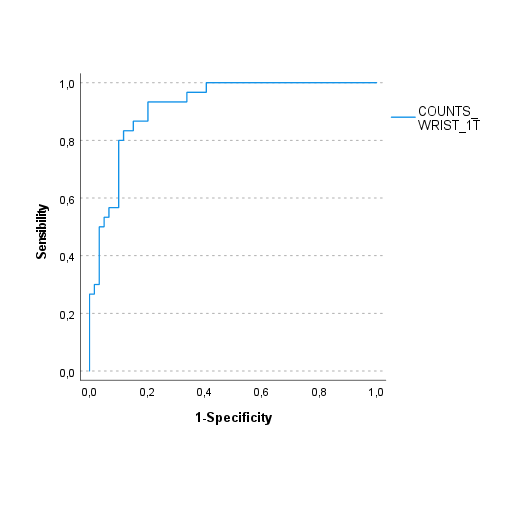

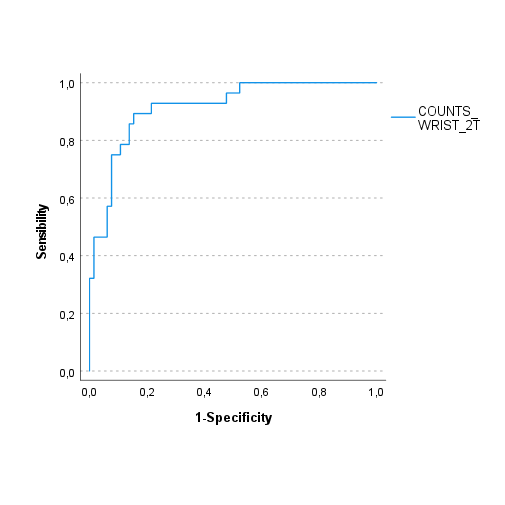

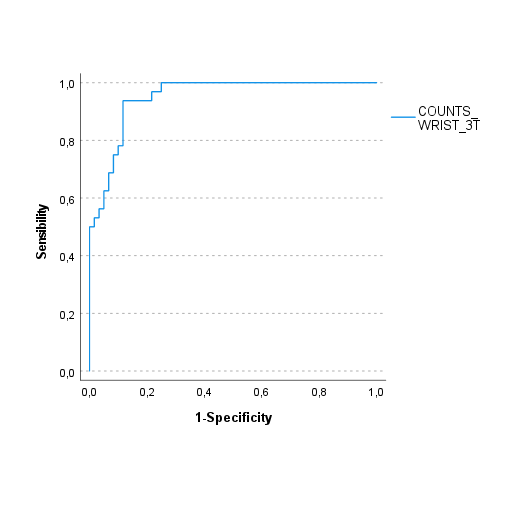

D	E	F

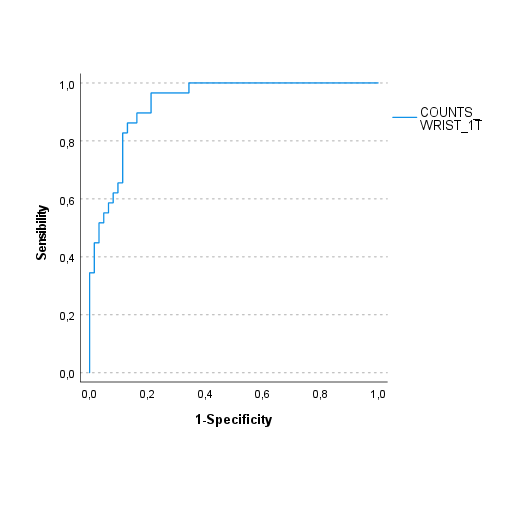

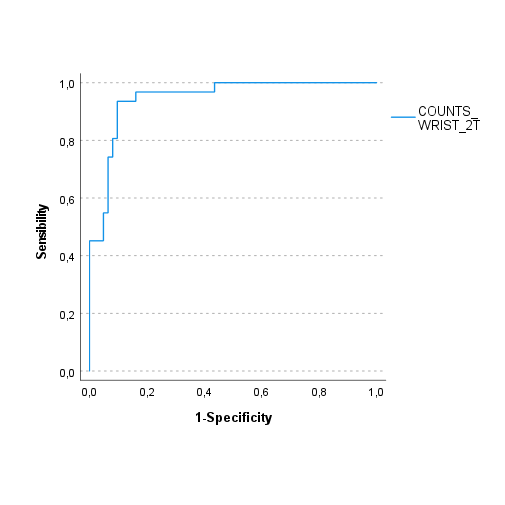

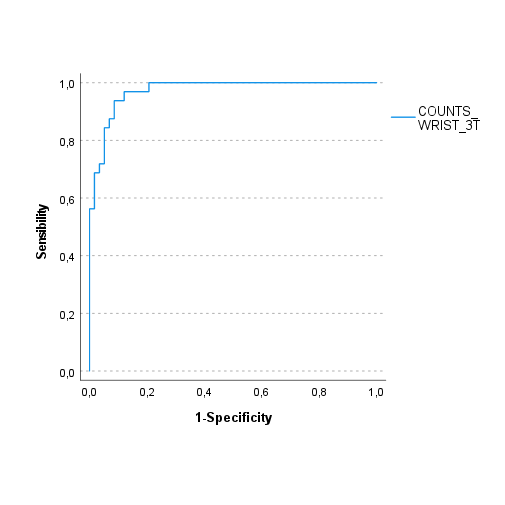

G	H	I

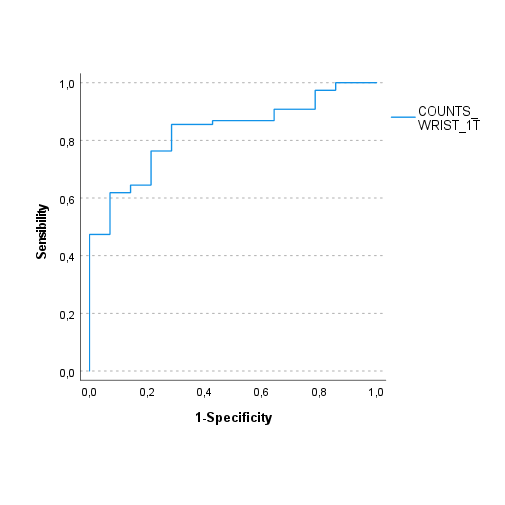

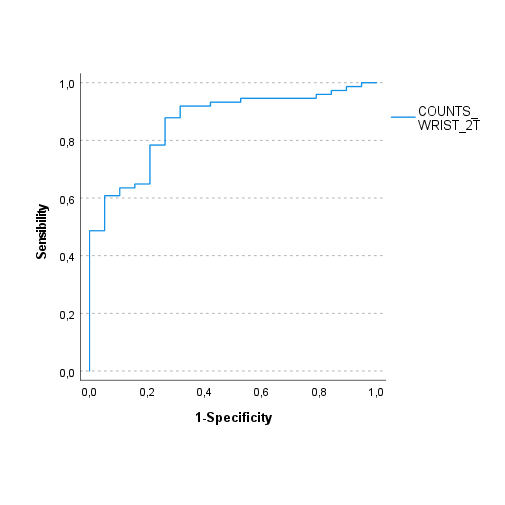

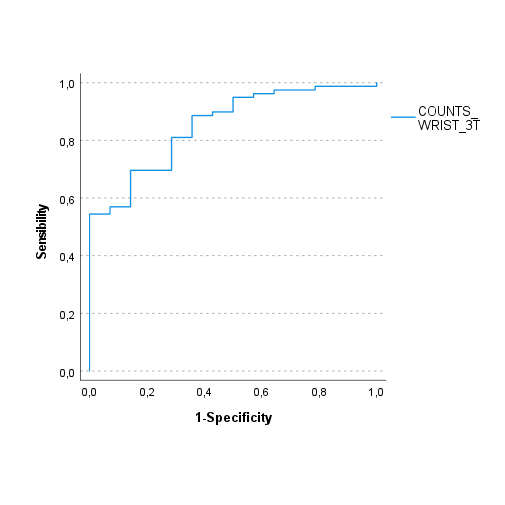

**Figure 5.** ROC curves for WRIST attachment in COUNTS units and MVPA cut-off points. A, B, C are ROC curves in 1T, 2T and 3T respectively using VO2R% criterion. D, E, F are ROC curves in 1T, 2T and 3T respectively using HRR% criterion. G, H, I are ROC curves in 1T, 2T and 3T respectively using MET criterion.
