## Supplemental Figure 10 for "Validation of Accelerometry Cut-off Points to Categorize Physical Activity Intensity Along the Pregnancy"

A	B	C

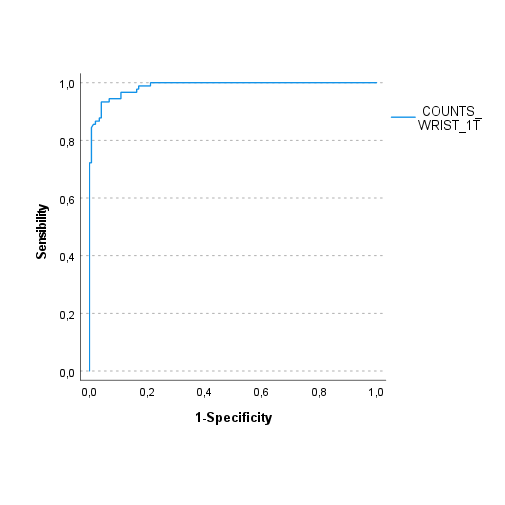

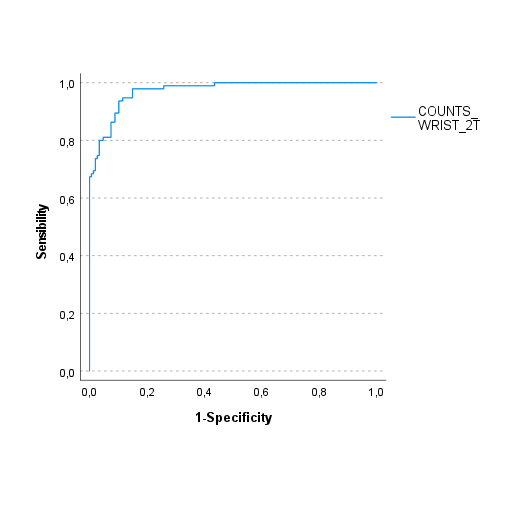

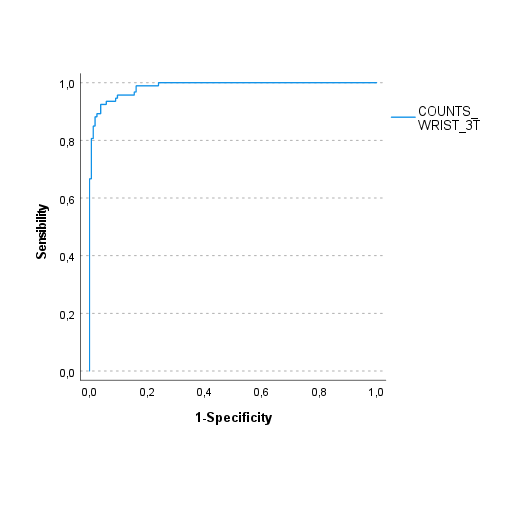

D	E	F

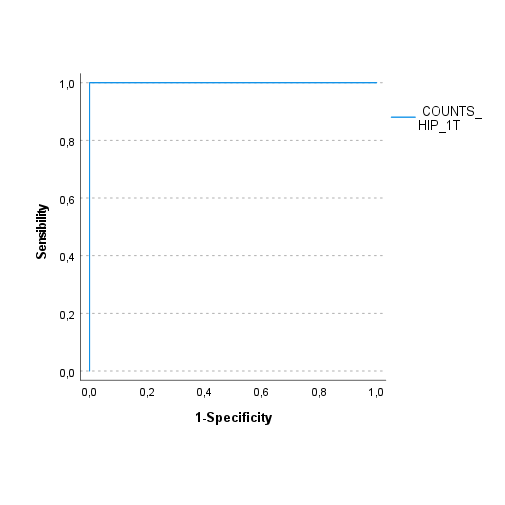

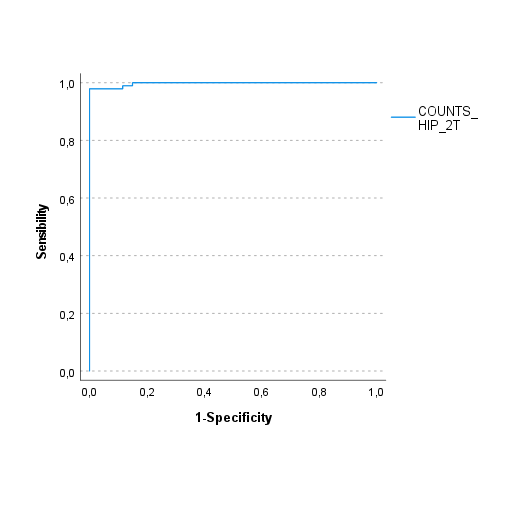

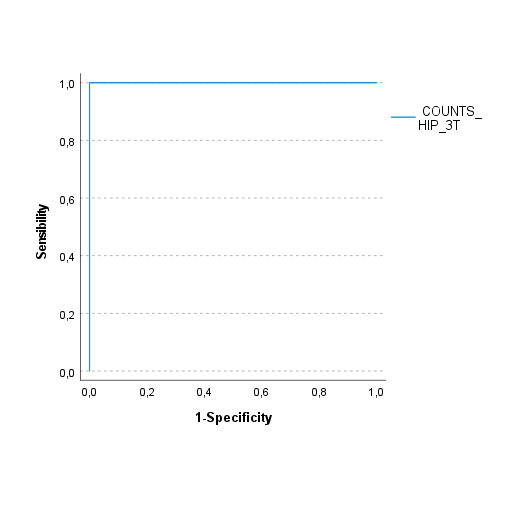

Figure *: ROC curves for WRIST and HIP COUNTS LPA cut-off points

A,B,C: ROC curves for WRIST in 1T,2T and 3T respectively using direct observation criteria.

D,E,F: ROC curves for HIP in 1T,2T and 3T respectively using observation criteria.
