## Supplemental Table 1 for "Validation of Accelerometry Cut-off Points to Categorize Physical Activity Intensity Along the Pregnancy"

**Table 1: intensity load in CPE-T along pregnancy**

| **Variable** |  |  | **1T** | | |  | **2T** | | |  | **3T** | | |
| --- | --- | --- | --- | --- | --- | --- | --- | --- | --- | --- | --- | --- | --- |
|  |  |  | Mean | SD | |  | Mean | SD | |  | Mean | SD | |
| **Speed 1** | **Load** | *(speed & slope)* | 2.0 km/h at 1% | | | | | | | | | | |
|  | **VO_2_** | *(L/min)* | 0.662^a^ | ± | 0.117 |  | 0.659^a^ | ± | 0.102 |  | 0.734^b^ | ± | 0.107 |
|  | **VO_2_** | *(ml/kg/min)* | 9.601 | ± | 1.217 |  | 9.271 | ± | 1.018 |  | 9.375 | ± | 0.952 |
|  | **%VO_2max_** | *(%)* | 29.2 | ± | 6.4 |  | 29.4 | ± | 6.1 |  | 30.5 | ± | 6.5 |
|  | **VO2R** | *(L/min)* | 0.358^a^ | ± | 0.09 |  | 0.353^a^ | ± | 0.076 |  | 0.401^b^ | ± | 0.805 |
|  | **%VO2R** | *(%)* | 18.3 | ± | 5.3 |  | 18.3 | ± | 5.0 |  | 19.4 | ± | 5.1 |
|  | **METS** |  | 3.0 | ± | 0.4 |  | 2.9 | ± | 0.3 |  | 3.0 | ± | 0.3 |
|  | **RPE** | *(Borg 0-10)* | 1.0 |  |  |  | 1.0 |  |  |  | 1.0 |  |  |
|  | **HR** | *(beats/min)* | 96.5^a^ | ± | 13.4 |  | 98.4^a^ | ± | 12 |  | 107.5^b^ | ± | 10.6 |
|  | **%HR_max_** | *(%)* | 52.2^a^ | ± | 7.1 |  | 53.3^a^ | ± | 6.4 |  | 58.2^b^ | ± | 5.6 |
|  | **HRR** | *(beats/min)* | 16.0 | ± | 6.6 |  | 17.5 | ± | 6.6 |  | 16.3 | ± | 6.0 |
|  | **%HRR** | *(%)* | 15.3 | ± | 6.4 |  | 17.0 | ± | 6.3 |  | 17.5 | ± | 6.5 |
| **Speed 2** | **Load** | *(speed & slope)* | 4.0 km/h at 1% | | | | | | | | | | |
|  | **VO_2_** | *(L/min)* | 0.896^a^ | ± | 0.157 |  | 0.910^a^ | ± | 0.153 |  | 1.026^b^ | ± | 0.171 |
|  | **VO_2_** | *(ml/kg/min)* | 13.018 | ± | 1.346 |  | 12.741 | ± | 1.284 |  | 13.018 | ± | 1.041 |
|  | **%VO_2max_** | *(%)* | 39.3^a^ | ± | 7.2 |  | 40.5^a,b^ | ± | 7.6 |  | 42.4^b^ | ± | 8.9 |
|  | **VO2R** | *(L/min)* | 0.593^a^ | ± | 0.133 |  | 0.604^a^ | ± | 0.123 |  | 0.693^b^ | ± | 0.144 |
|  | **%VO2R** | *(%)* | 30.0^a^ | ± | 6.6 |  | 31.2^a,b^ | ± | 7.0 |  | 33.4^b^ | ± | 8.3 |
|  | **METS** |  | 4.1 | ± | 0.4 |  | 4.0 | ± | 0.4 |  | 4.1 | ± | 0.3 |
|  | **RPE** | *(Borg 0-10)* | 2.0 |  |  |  | 2.0 |  |  |  | 3.0 |  |  |
|  | **HR** | *(beats/min)* | 107.6^a^ | ± | 13.6 |  | 109.7^a^ | ± | 13 |  | 119.6^b^ | ± | 11.8 |
|  | **%HR_max_** | *(%)* | 58.3^a^ | ± | 7.2 |  | 59.4^a^ | ± | 7.0 |  | 64.7^b^ | ± | 6.3 |
|  | **HRR** | *(beats/min)* | 27.1 | ± | 7.7 |  | 28.8 | ± | 8.4 |  | 28.4 | ± | 6.9 |
|  | **%HRR** | *(%)* | 26.2^a^ | ± | 7.1 |  | 28.0^b^ | ± | 8.1 |  | 30.7^c^ | ± | 8.2 |
| **Speed 3** | **Load** | *(speed & slope)* | 6.0 km/h at 1% | | | | | | | | | | |
|  | **VO_2_** | *(L/min)* | 1.404^a^ | ± | 0.264 |  | 1.433^a^ | ± | 0.268 |  | 1.636^b^ | ± | 0.281 |
|  | **VO_2_** | *(ml/kg/min)* | 20.270 | ± | 2.040 |  | 20.111 | ± | 2.057 |  | 20.864 | ± | 1.767 |
|  | **VO_2max%_** | *(%)* | 61.6^a^ | ± | 12.0 |  | 63.8^a,b^ | ± | 13.6 |  | 67.7^b^ | ± | 15.2 |
|  | **VO2R** | *(L/min)* | 1.101^a^ | ± | 0.241 |  | 1.127^a^ | ± | 0.239 |  | 1.303^b^ | ± | 0.261 |
|  | **VO2R%** | *(%)* | 55.9^a^ | ± | 12.9 |  | 58.4^a,b^ | ± | 14.7 |  | 62.9^b^ | ± | 16.6 |
|  | **METs** |  | 6.4 | ± | 0.6 |  | 6.4 | ± | 0.7 |  | 6.6 | ± | 0.6 |
|  | **RPE** | *(Borg 0-10)* | 4.0 |  |  |  | 4.0 |  |  |  | 5.0 |  |  |
|  | **HR** | *(beats/min)* | 137.5^a^ | ± | 15.5 |  | 140.6^a^ | ± | 18 |  | 149.8^b^ | ± | 16.1 |
|  | **HR_max%_** | *(%)* | 74.5^a^ | ± | 8.6 |  | 76.1^a^ | ± | 10 |  | 81.1^b^ | ± | 9.0 |
|  | **HRR** | *(beats/min)* | 56.9 | ± | 12.9 |  | 59.7 | ± | 11.1 |  | 58.6 | ± | 13.9 |
|  | **%HRR** | *(%)* | 55.1^a^ | ± | 12.6 |  | 58.6^a^ | ± | 14.5 |  | 63.3^b^ | ± | 16.1 |

Values are mean and standard deviation except in Borg scale, where data are in median; n=31. 1T, first trimester; 2T, second trimester; 3T, third trimester; VO_2,_ oxygen consumption_;_ VO_2max_%, maximum oxygen consumption percentage; VO_2R_, oxygen consumption reserve; VO_2R_%, oxygen consumption reserve percentage; METs, metabolic equivalents; RPE, rating of perceived exertion ; HR, heart rate; HR_max_%, maximal heart rate percentage; HRR, heart rate reserve; HRR% heart rate reserve percentage; a, b, c letters indicate significant differences between trimesters (*P*<0.05).
