## Supplemental Table 2 for "Validation of Accelerometry Cut-off Points to Categorize Physical Activity Intensity Along the Pregnancy"

**Table 2. Counts, MIMS and ENMO data registered at each speed during pregnancy for both ACLs placements.**

| **Variable** | | **1T** | | |  | **2T** | | |  | **3T** | | |
| --- | --- | --- | --- | --- | --- | --- | --- | --- | --- | --- | --- | --- |
|  |  | Mean | SD | |  | Mean | SD | |  | Mean | SD | |
| **Speed 1** | **Load** | 2.0 km/h at 1% | | | | | | | | | | |
|  | **Counts VM**_wrist_ | 2352.3 | ± | 1043.7 |  | 2523.8 | ±± | 995.89 |  | 2371.68 | ±± | 923.7 |
|  | **Counts VM**hip | 1828.9 | ± | 521.3 |  | 1854.2 | ± | 716.9 |  | 1772.4 | ± | 448.8 |
| **Speed 2** | **Load** | 4.0 km/h at 1% | | | | | | | | | | |
|  | **Counts VM**_wrist_ | 4120.9 | ± | 1675.6 |  | 4064.28 | ± | 1367.8 |  | 4475.6 | ± | 1469.3 |
|  | **Counts VM**hip | 3447.5 | ± | 630 |  | 3422.7 | ± | 701.3 |  | 3454.6 | ± | 621.4 |
| **Speed 3** | **Load** | 6.0 km/h at 1% | | | | | | | | | | |
|  | **Counts VM**wrist | 7613.2^a^ | ± | 2360.2 |  | 8511^b^ | ± | 3051.1 |  | 9583.8^c^ | ± | 3654.8 |
|  | **Counts VM**hip | 5793.1 | ± | 853.5 |  | 5646.2 | ± | 823.0 |  | 5624.5 | ± | 825.2 |
| **Speed 1** | **Load** | 2.0 km/h at 1% | | | | | | | | | | |
|  | **MIMS _wrist_** | 19.11 | ± | 5.83 |  | 16.72 | ± | 4.10 |  | 16.18 | ± | 4.09 |
|  | **MIMS** hip | 13.25 | ± | 2.00 |  | 12.98 | ± | 1.92 |  | 12.53 | ± | 1.20 |
| **Speed 2** | **Load** | 4.0 km/h at 1% | | | | | | | | | | |
|  | **MIMS _wrist_** | 22.05 | ± | 5.71 |  | 21.78 | ± | 4.77 |  | 22.91 | ± | 5.31 |
|  | **MIMS** hip | 23.91 | ± | 3.15 |  | 23.37 | ± | 2.93 |  | 22.61 | ± | 1.88 |
| **Speed 3** | **Load** | 6.0 km/h at 1% | | | | | | | | | | |
|  | **MIMS _wrist_** | 33.81^a^ | ± | 7.18 |  | 36.87^b^ | ± | 12.05 |  | 38.44^c^ | ± | 10.63 |
|  | **MIMS** hip | 41.84 | ± | 5.19 |  | 41.31 | ± | 5.49 |  | 40.86 | ± | 4.56 |
| **Speed 1** | **Load** | 2.0 km/h at 1% | | | | | | | | | | |
|  | **ENMO _wrist_** | 59.897 | ± | 16.85 |  | 59.915 | ± | 15.968 |  | 62.09 | ± | 15.625 |
|  | **ENMO** hip | 49.541^a^ | ± | 14.41 |  | 40.265^b^ | ± | 13.509 |  | 32.363^c^ | ± | 8.935 |
| **Speed 2** | **Load** | 4.0 km/h at 1% | | | | | | | | | | |
|  | **ENMO _wrist_** | 120.094 | ± | 36.511 |  | 115.501 | ± | 31.592 |  | 122.957 | ± | 35.541 |
|  | **ENMO** hip | 110.516^a^ | ± | 19.931 |  | 101.898^b^ | ± | 20.111 |  | 91.04^c^ | ± | 12.589 |
| **Speed 3** | **Load** | 6.0 km/h at 1% | | | | | | | | | | |
|  | **ENMO _wrist_** | 257.143^a^ | ± | 71.89 |  | 283.44^b^ | ± | 110.9 |  | 327.435^c^ | ± | 152.4 |
|  | **ENMO** hip | 219.91^a^ | ± | 33.796 |  | 214.963^a,b^ | ± | 37.188 |  | 203.408^b^ | ± | 37.073 |

Values are mean and standard deviation (SD); n=31. 1T, first trimester; 2T, second trimester; 3T, third trimester; a, b, c letters indicate significant differences between trimesters (*P*<0.05).
